# Association of cultural participation with morbidity and mortality in the Welsh Population: An observational study

**DOI:** 10.1101/2023.08.24.23294550

**Authors:** Daniel Peter Jones, Lisa Hurt, Rhian Daniel

## Abstract

**Background:** Clinical interventions based on cultural participation have consistently demonstrated improved patient outcomes. At the population level, prospective cohort studies show a protective association between cultural participation and all-cause mortality. However, the latter is limited by a narrow choice of methods and outcomes. We sought to address this by using multiple methods to investigate the effects of cultural participation on both morbidity and mortality.

**Methods:** First, we conducted multivariable linear and logistic regression models using the National Survey of Wales 2015-2019. This featured attendance at cultural events/activities as an exposure and self-reported health (SRH), as well as specific health conditions, as an outcome with sequential adjustment for confounders related to demographics, socio-economic position (SEP), and lifestyle behaviours. To complement this, we conducted a multivariable panel regression model using cultural expenditure data from English and Welsh local authorities and life expectancy data from the Office for National Statistics.

**Results:** When adjusting for age, sex, SEP, and lifestyle behaviours, we found that overall cultural participation had a small positive association with SRH (4.8% improvement in SRH score, 95% confidence intervals [4.0-5.6%], p<0.0001, n=15374) and lower likelihood of cardiovascular disease, and anxiety/depression. Similar, albeit smaller, positive associations were seen between SRH and individual cultural activities, including library, museum, and heritage site visits. However, we did not identify any association between local authority cultural expenditure and local life expectancy.

**Conclusion:** We identified new evidence for a positive association between cultural participation and morbidity. However, additional evidence will be required to better establish causal inference.

**What is already known on this topic?:** Several observational studies show that cultural participation is associated with reduced mortality and clinical trials also support the use of cultural interventions.

**What this study adds?:** This study provides evidence for beneficial associations between cultural participation, self-reported health, and certain chronic conditions even when taking account of known confounders.

**How this study might affect research, practice or policy?:** This study further emphasises the importance of arts and culture for health and the need for further research in this area of epidemiology.

## BACKGROUND

Culture and the arts form part of everyday life. Definitions vary with a wide range of views held on the subject, for example Kant’s aesthetics [1] or Bourdieu’s cultural capital[2]. The World Health Organisation (WHO) Europe programme for cultural insights into health broadly defines cultural activities as having three key characteristics: being valuable in their own right; providing imaginative experiences; and provoking an emotional response [3]. Within the UK, the Welsh Government uses constituent examples, specifically including attendance/participation at arts events (film, live music, theatre, dance, storytelling), museums, heritage sites and libraries/archives [4].

The potential for culture to influence health has attracted wide-spread recent interest. For example, there is now a UK-wide, cross-sectoral alliance in the area [5] and WHO Europe has commissioned a programme on the arts and health [3]. The UK Department for Culture, Media and Sport has also published an in-depth report on the arts and health [6]. These reports provide strong evidence for individual- and group-level clinical/non-clinical cultural interventions promoting health and well-being. Evidence from systematic reviews identifies that community art engagement reduces social isolation and loneliness [7], dance class programmes reduce fall risk in older adults [8] and that music therapy can improve mental health behaviours in children and adolescents [9]. The mechanisms are likely due to a combination of social engagement, physical activity, access to green space, health literacy, sensory activation, cognitive stimulation, and emotional evocation [10]. Economic evaluations have found such programmes to generally be cost-effective or saving[11]. For example, the ‘Arts on Prescription’ programme in England is estimated to net-save the NHS £216 per prescription through reduced healthcare utilisation [12].

Epidemiologically, prospective cohort studies have found that frequent cultural participation is associated with reduced all-cause mortality (as much as 20% lower) when compared to those who never participate [13-14]. However, such observational work is subject to potential bias from unknown confounders and reverse causation.

In this study, we sought to further explore associations of cultural participation with morbidity and mortality in the general population of a high-income country We used multiple methods as part of a triangulation approach to help support causal inference by covering respective methodological flaws [15].

## METHODS

### I. INDIVIDUAL-LEVEL ANALYSIS

#### SAMPLE

This analysis used the National Survey for Wales (NSW), dated 2016-2020 (inclusive) [16]. The NSW began in 2016 and data from March 2020 onwards was not used to avoid the potential impact of the Covid-19 pandemic.

The NSW is a representative^i^ survey of the Welsh general population, aged over 16 years, conducted by the Office for National Statistics (ONS) throughout the year, via face-to-face interview, with annual reports [17]. It provides information on a range of cultural participation measures and recent self-reported health in general as well as information on multiple potential confounding factors [18-19]. All data are publicly available.

Random sampling of Welsh households took place with oversampling of certain local authorities and ensure proportionate geographical coverage across Wales. Eligible households received notification by post in advance and are then contacted by phone with non-contacts being re-contacted up to three times. In 2018/19, the survey had a response rate of 48% with 34% refusing, 6% not contactable and 12% being ineligible.

All persons aged over 16 years in a selected household were eligible person in each household selected at random to participate. Participants received shopping vouchers as a financial incentive for completion. Typically, each annual report featured 12000 participants. Further technical details are publicly available [20].

#### MEASURES

Cultural participation (exposure): Participants provided information on a range of recent activities. Participants reported frequency in the last 12 months of library attendance, arts event^ii^ attendance, participation in a cultural activity^iii^, heritage site^iv^ visit, and museum attendance. Attendance information was reported in a binary manner (yes/no) where yes represented undertaking at least one of the listed activities in the last 12 months. For individual activities, attendance was also reported as a binary outcome (yes/no) where yes represented undertaking the activity at least once in the last 12 months.

Self-reported health (primary outcome): Participants were asked to report their “health in general” with five options presented to choose from (very bad/bad/fair/good/very good). These were coded 0, 1, 2, 3 and 4 respectively for statistical analysis.

Long-term illness (secondary outcome): Participants who reported chronic illness (an illness lasting longer than 12 months) were asked to specify which one. These responses were then coded according to ‘International Classification of Diseases 10’ chapter [21]. The chapter codes of interest are V and IX (“Mental and behavioural disorders” & “Diseases of the circulatory system” respectively).

Co-variates: These included age, sex, ethnic group, employment status (employed/unemployed/retired), educational attainment (measured by respective National Vocational Qualification level), and local authority. Further covariates included illness/disability lasting longer than 12 months that limited daily activity at least somewhat, exercise/sport participation in last 4 weeks, smoking status, and alcohol drinking status.

#### STATISTICS

All analyses were performed using the latest R statistical package 4.1.0 [22]. A directed acyclic graph [23] showing the relationships between the exposure, mediators, confounders, and outcome is shown in Figure 1 overleaf. Power calculations were not performed given that relevant effects have been detected by comparable studies with smaller samples from the same data sources (Gott, 2020). In contrast, this analysis used the maximally available number of participants from several combined surveys (n ∼ 48000) although this number was smaller for certain analyses, due to incomplete data. When planning our analyses, we used small workshops involving professional stakeholders and members of the public recruited via Health and Care Research Wales.

**Figure 1.**
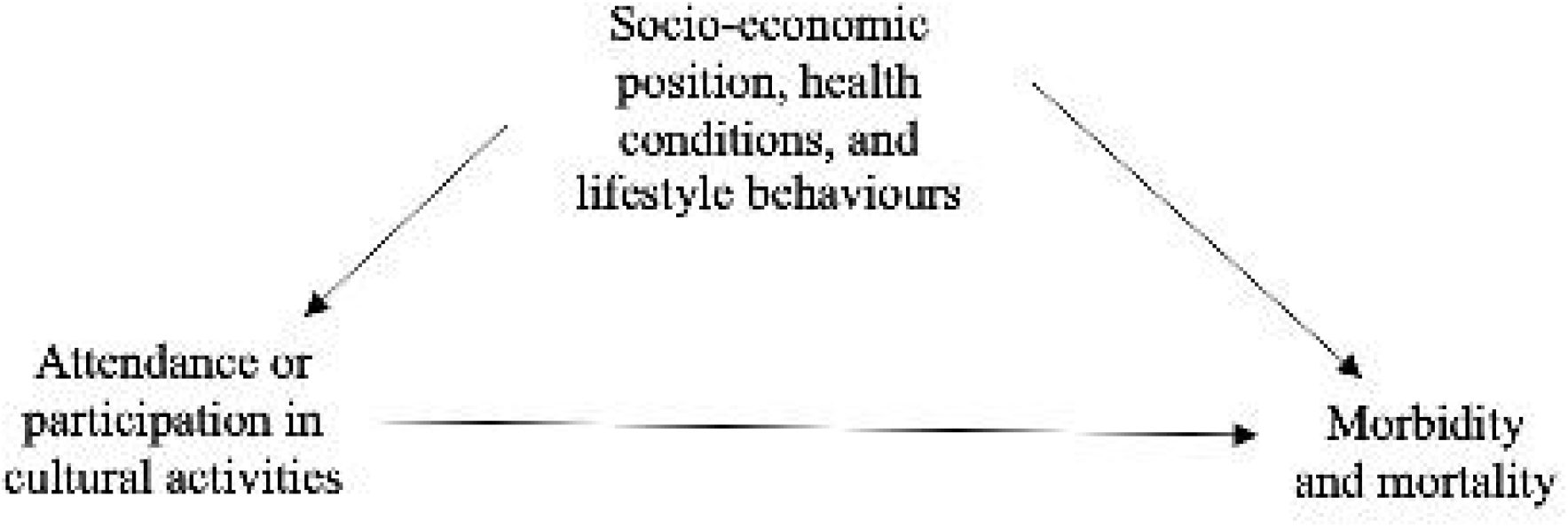
A directed acyclic graph demonstrating the pathway between cultural participation, confounders and health outcomes

Chi^2^ analyses were performed to assess relations between basic demographic factors and cultural participation. Multivariable linear regression models were used with self-reported health (scored by ordinal ranking) as a primary outcome and library attendance, arts event attendance, cultural activity participation and museum attendance as a composite exposure. This was followed by analyses with each specific cultural activity. Secondary outcomes were anxiety/depression and cardiovascular disease as both are partly linked to psycho-social causes [24-25].

A sequential adjustment approach was used. In the first set of models, results were only adjusted for age, sex, ethnicity, local authority, and survey year. In the second set of models, additional adjustment took place for individual socio-economic position (SEP) markers (educational attainment and employment status) and chronic illness. In a final set of models, adjustment also took place for lifestyle behaviours (smoking, alcohol, and exercise). Individuals with missing data were excluded.

### II. AREA-LEVEL ANALYSIS

#### SAMPLE

Publicly available data were extracted for the 22 Welsh local authorities (each ranging from 60,000 people to 370,000) and the 58 English unitary authorities (distributed throughout the country, excluding Greater London, with populations ranging from 40,000 to 550,000). Data was collected from between 2011 and 2019.

#### MEASURES

Local authority culture spending (exposure): All Welsh local government net expenditure in Wales is available via the Welsh Government [26]. All English local government data for unitary authorities is available via the Department for Levelling Up, Housing, and Communities [27]. All spending data was adjusted for inflation with a 2019 baseline using a Gross Domestic Product deflator [28] and for population using ONS estimates.

Mortality data (outcome): The ONS regularly publishes life tables, including three year rolling averages of life expectancy and healthy life expectancy, using routine demographic information [29].

#### STATISTICS

A fixed-effects multivariable panel regression model was used with male and female life expectancy and healthy life expectancy between 2010 and 2019 used as outcomes and each local area forming a unit. Annual combined per capita library, cultural and heritage local authority spending was used as an exposure allowing for a lag of 3 years. Panel regression theoretically accounts for any confounders that are time invariant across local authorities. Nonetheless, total local authority spending was adjusted for.

### III. SENSITIVITY ANALYSES

To address uncertainty, a number of pre-specified sensitivity analyses were conducted. For the individual analyses, we conducted two further analyses involving people living in the most deprived and least deprived areas respectively to assess for possible effect modification.

For the area-level analysis, we performed an additional analysis using life expectancy and healthy expectancy at 65 as an outcome. We also varied the time lag for any effect up to a maximum lag of 5 years. Finally, we performed separate analyses for Welsh and English local authorities.

## RESULTS

### SAMPLE CHARACTERISTICS

A representative sample of 46,178 participants was included from the 2016-2020 NSWs with demographic details shown in table 1 overleaf. 15,716 participants had complete data for all analyses (for example some questions were only asked to a smaller random subsets).

**TABLE 1.**
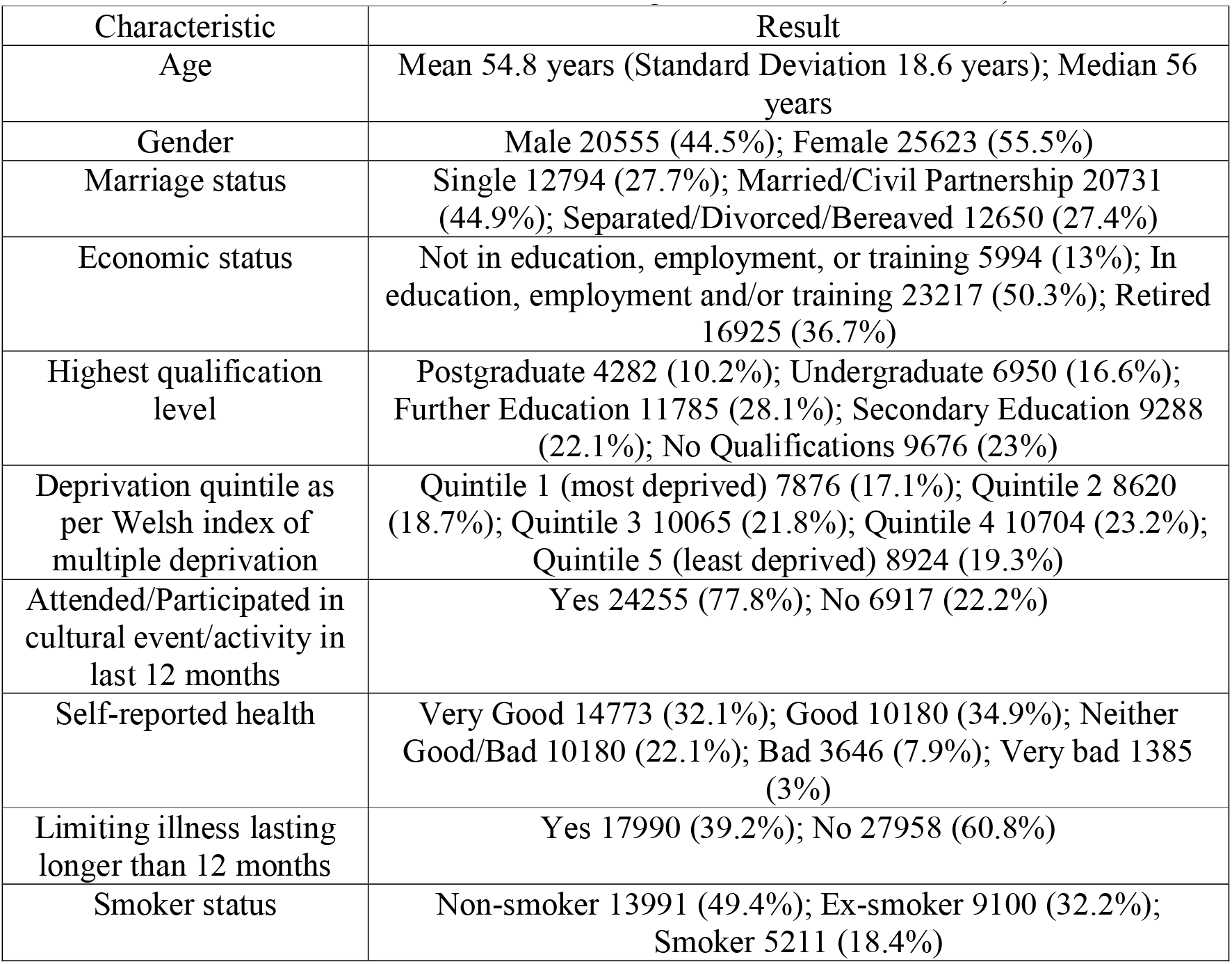
A table illustrating the demographic details from the National Survey for Wales sample (Total n=46189 however data are missing for certain characteristics)

In total, 24,255 (77.8%) of participants had attended/participated in 3 or more cultural activities or events in the last 12 months whilst 6917 (22.2%) had not. Cultural participation was negatively associated with low socio-economic position and/or long-term illness or disability, as confirmed by Chi^2^ analyses shown in Table 2. Data to measure further aspects of inequality, such as racial inequalities, were not available.

**TABLE 2.** A table demonstrating the associations between certain demographic characteristics and cultural participation.

| Characteristic | Attended/Participated in Cultural Event/Activity in last 12 months |  | Chi<sup>2</sup> Result |
| --- | --- | --- | --- |
|  | Yes | No |  |
| Most deprived quintile | 4308 (68.1%) | 2020 (31.9%) | Chisq = 689.1, df = 4, p-value <0.0001 |
| Least deprived quintile | 5026 (86.5%) | 785 (13.5%) |  |
| Postgraduate qualification | 2657 (93.9%) | 173 (6.1%) | Chisq = 3919, df = 4, p-value<0.0001 |
| No qualification | 3463 (52.1%) | 3183 (47.9%) |  |
| In employment, education and/or training | 13756 (87.7%) | 1937 (12.3%) | Chisq = 1819.4, df = 2, p-value <0.0001 |
| Not in employment, education, or training | 2708 (64.4%) | 1500 (35.6%) |  |
| Limiting illness lasting longer than 12 months | 8465 (68.0%) | 3980 (32.0%) | Chisq = 1158.2, df = 1, p-value <0.0001 |
| No limiting illness lasting longer than 12 months | 15700 (84.3%) | 2904 (15.7%) |  |

### ASSOCIATION OF CULTURAL PARTICIPATION WITH MORBIDITY

We found a small, protective association between cultural participation and morbidity, shown in Table 3. Attending 3 or more cultural activities/events in the past 12 months was associated with an adjusted 4.8% improvement in SRH score (95% confidence intervals [CI] 4-5.6%, p<0.0001, n=15734).

**TABLE 3.** A table demonstrating the associations between cultural participation and self-reported health with staged adjustment for potential confounders.

| Exposure | Percentage Change in Self-Reported Health Score (95% Confidence Intervals) | P-value | Number of participants included |
| --- | --- | --- | --- |
| Cultural Participation 1 | +14.3% (13.6-15.0%) | <0.0001 | 31129 |
| Cultural Participation 2 | +6.3% (5.7-6.9%) | <0.0001 | 28170 |
| Cultural Participation 3 | +4.8% (4.0-5.6%) | <0.0001 | 15734 |

- 1- Adjusted for age and gender - 2- Additionally adjusted for socio-economic position and limiting illness - 3- Additionally adjusted for health behaviours

Similarly, as shown in Table 4, when fully adjusted for known confounders, individuals attending 3 or more cultural activities/events in the past 12 months were 28% less likely to report cardiovascular disease (Adjusted odds ratio [OR] 0.72, 95% CI 0.58-0.90, p<0.003, n=15716). Frequent cultural participation was also associated with a reduced likelihood of reporting anxiety or depression although the confidence intervals crossed the null in the most adjusted model (Adjusted OR 0.91, 95% CI 0.80-1.04, p=0.19, n=15716).

**TABLE 4.**
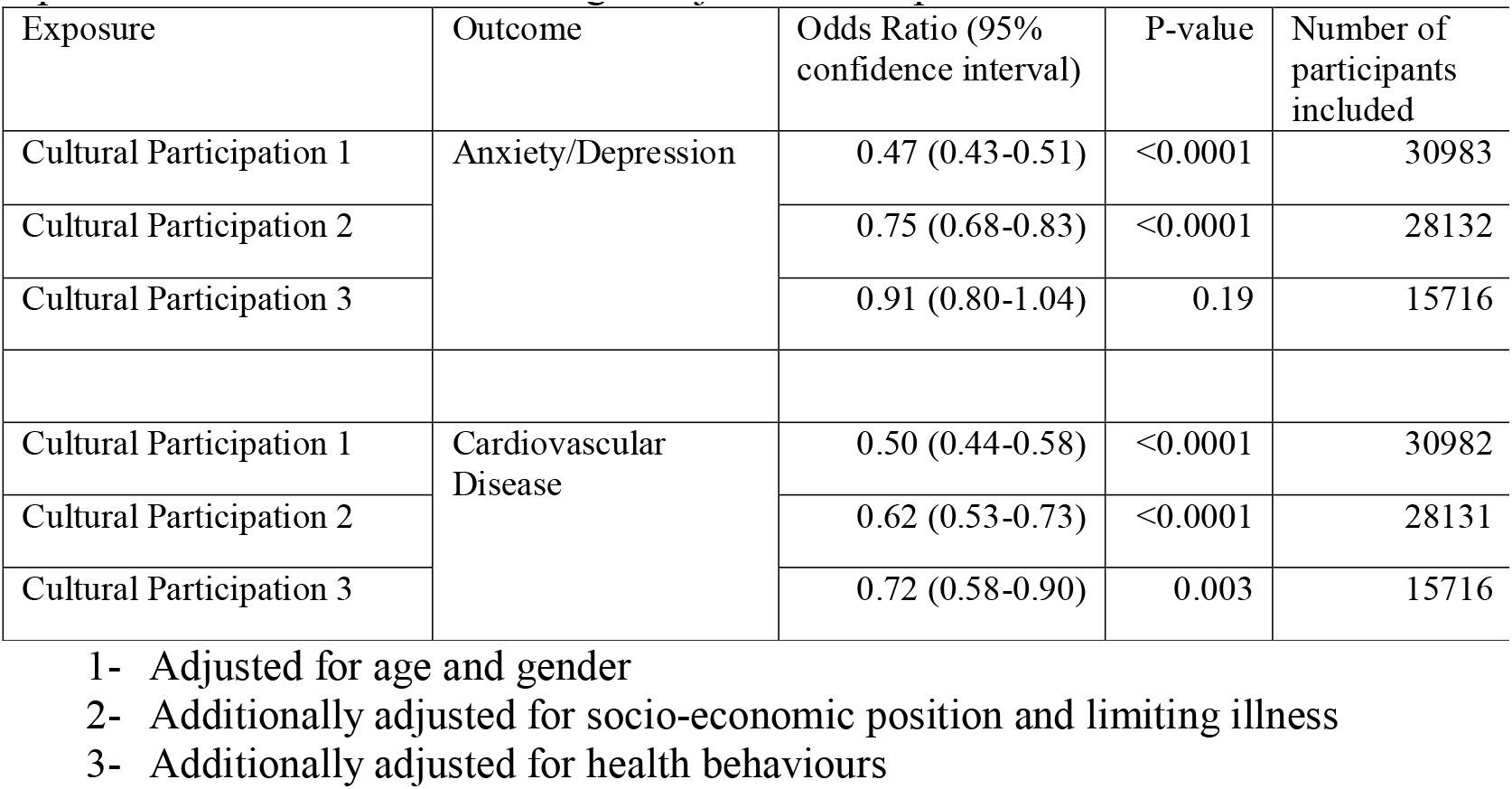
A table demonstrating the associations between cultural participation and specific health outcomes with staged adjustment for potential confounders.

Finally, we saw small, positive associations between specific cultural activities (heritage site attendance, museum attendance, and library/archive attendance) and SRH (See Table 5). In a sensitivity analysis, we found no evidence for effect modification by socio-economic position.

**TABLE 5.** A table demonstrating the associations between specific cultural activities/events and self-reported health with staged adjustment for potential confounders.

| Exposure | Percentage Change in Self-Reported Health Score (95% confidence interval) | P-value | Number of participants involved |
| --- | --- | --- | --- |
| Heritage 1 | +9.6% (9.0-10.2%) | <0.0001 | 27066 |
| Heritage 2 | +3.2% (2.6-3.7%) | <0.0001 | 24485 |
| Heritage 3 | +2.6% (1.7-3.5%) | <0.0001 | 13845 |
| Museum Visit 1 | +8.7% (8.1-9.4%) | <0.0001 | 24909 |
| Museum Visit 2 | +3.1% (2.5-3.7%) | <0.0001 | 22517 |
| Museum Visit 3 | +2.4% (1.5-3.2%) | <0.0001 | 10118 |
| Library/Archive 1 | +9.6% (9-10.2%) | <0.0001 | 27066 |
| Library/Archive 2 | +3.2% (2.6-3.7%) | <0.0001 | 24485 |
| Library/Archive 3 | +1.0% (0.4-1.5%) | 0.001 | 20196 |

- 1- Adjusted for age and gender - 2- Additionally adjusted for socio-economic position and limiting illness - 3- Additionally adjusted for health behaviours

### ASSOCIATION OF CULTURAL SPENDING WITH MORBIDITY & MORTALITY

We did not find any association between per capita expenditure on culture and life expectancy or healthy life expectancy in English and Welsh local authorities when adjusted for total expenditure. A lack of association was consistent for both male and female life expectancies at both birth and age 65. In sensitivity analyses, we used a range of lag times for an effect to take place but still failed to identify any association.

## DISCUSSION

In this study, we used a range of methods and observational data to further explore links between cultural participation and morbidity and mortality. Our results demonstrate that, in the Welsh population, there is a positive association between cultural participation and self-reported health across a range of different activities even when accounting for a number of known confounders. Similarly, we found that cultural participation was associated with a reduced likelihood of reporting cardiovascular disease, anxiety, or depression.

We sought to further investigate this with different data and methods, namely local authority expenditure on culture in England and Wales. However, we were not able to identify any associations between this and life expectancy or healthy life expectancy.

### OUR RESULTS IN CONTEXT

Our results complement existing work in this area building on the results from several large, prospective cohort studies conducted in recent years in Japan, the United States, England, Scotland, Norway, Sweden, and Finland[13, 30-34]. Those studies measured cultural participation at a baseline in cohorts and followed up participants through linked death registries over several years. Cultural participation was consistently associated with a 10-20% reduction in all-cause mortality, even when adjusted for important confounders, and displayed a dose-response phenomenon. Our work now demonstrates a further association between cultural participation and self-reported health.

Our work is also consistent with analogous research, such as Scottish Government analysis of cultural participation and health links in the Scottish Household Survey [35]. In recent work, Alexiou *et al*. established that reductions in per capita local authority spending in England were associated with small but significant reductions in male and female life expectancy at birth and at 65 (roughly a month reduction for every £100 of per capita spend lost)[36]. However, this study investigated the totality of local authority spending, including social care and key determinants of health, such as housing assistance and financial support. Given the relatively small population-level effect identified in this scenario, any effect of reduced cultural spending may be too small detect even with the timescale and local authority numbers we used.

### IMPLICATIONS

Our study presents several implications for public health and policy. Firstly, it re-affirms the social patterning, by socio-economic position and disability, of cultural participation in high-income countries [4]. Further work is needed to reduce these inequalities and, as our stakeholders repeatedly emphasised, to make both culture and the measurement of culture more inclusive and diverse.

Secondly, our study established a positive association between cultural participation and self-reported health that remained consistent over time and across activity types. Similarly, it found beneficial associations between cultural participation and certain health conditions, including cardiovascular disease and mental illness. These associations remained consistent even when adjusted for age, sex, socio-economic indicators, long-term illness/disability, and unhealthy lifestyle behaviours highlighting the potential of cultural participation as a universal health benefit for populations [6] and the potential enabling role of health for cultural participation.

Finally, our study established that changes in cultural spending do not necessarily translate into measurable effects on population health. Evidently, it is important to focus on cultural participation itself as a mechanism of action, rather than just investment, to be realistic about the speed and scale of health benefits it can deliver, and to appreciate the wider benefits of cultural participation, such as economic stimulus and increased community cohesion [37].

### STRENGTHS AND LIMITATIONS

The strengths of our study included the use of enumerated data from publicly available, high-quality sources that featured a robust definition of cultural participation, comparable to that used in wider literature. This data allowed for sequential adjustment of several known confounders of cultural participation and health and supplemented with several sensitivity analyses to reduce uncertainty. It also featured reliable measures for health outcomes, allowing us to generate novel findings on morbidity. Finally, we used additional methods, namely ecological data, and analyses, as part of a triangulation approach [15], to further reduce uncertainty around causal inference.

A limitation of this observational data was its limited scope in both time and place, focussing on samples of the Welsh population over the course of 5 years. This reflected a lack of other population surveys in the UK (and beyond) that regularly ask about both health and cultural participation, using robust and reliable measures. Furthermore, the NSW has only recently introduced such measures and we limited the data to the time period before the Covid-19 pandemic, given the profound effects that this had on both cultural participation and population health. Nevertheless, in spite of a limited sample with incomplete data for a number of subjects, our study was able to detect an important association with adequate precision as demonstrated by the relevant confidence intervals. Our observational data was also limited by possible reverse causation and the possibility of bias from confounding. Regarding the former, we conducted an additional analysis that did measure changes in culture spending and life expectancy measures over time in a panel regression model. We sought to deal with the latter by adjusting for known potential confounders, including chromic health conditions, using multivariable regression in a sequential manner, results remained positive although residual confounding remains possible. We also conducted a series of sensitivity analyses that included isolating specific activities and investigating for effect modification with results continuing to support our conclusions.

Finally, there were limits apparent in our ecological analysis both in terms of the exposure measure and the number of contributing units in the panel regression. Data on culture is generally limited and, although less than ideal, the culture spending data we used was nonetheless available on an annual basis and at a local authority level. The limits in unit numbers arose from both measures only being available at a unitary authority level and the time period used deliberately avoiding overlap with the 2007-8 financial crisis and the Covid-19 pandemic. Accordingly, we additionally used English local authority data and varied time lags for effects in order to increase statistical power and although our final model failed to detect any associations, it did have moderately good precision suggesting small effect size or a lack of any effect was the most likely explanation of the results.

## CONCLUSIONS

In conclusion, our study has established an association between cultural participation and morbidity in the Welsh population. Higher cultural participation was associated with improved self-reported health and reduced likelihood of reporting cardiovascular disease, anxiety, and depression. These findings offer several important public health implications and strengthen the case for improving cultural participation.

## Data Availability

All data produced are available online at

https://gov.wales/national-survey-wales

https://www.gov.uk/government/organisations/department-for-levelling-up-housing-and-communities

https://www.ons.gov.uk/peoplepopulationandcommunity/healthandsocialcare/healthandlifeexpectancies

https://statswales.gov.wales/Catalogue/Local-Government/Finance/Revenue/Outturn

## ACKNOWLEDGEMENTS

We are grateful to the participants in the National Survey for Wales, and to the public representatives and professional stakeholders who we involved in the planning stages of this study.

## Footnotes

i Demographic sample statistics from 2018-19 include a male: female ratio of 49: 51, 50% married, 42% aged 16-44, 33% aged 45-64, 25% 65+, 92% white Welsh/British, 2-11% in each Welsh local authority area

ii Examples include play/drama/pantomime/musical; Live music event; Opera/classical music performance; Film at an arts centre; Carnival/street arts/arts festival; Exhibition or collection of art, craft, photography, or sculpture; Event including video art or electronic art; Event connected with books or writing; Circus (not involving animals); or dance performance

iii Examples include music of any kind; Drama or theatrical activity; Dance activity; Film or video making, or photography; Visual arts and crafts; Creative writing; Creating or making artwork or animation using digital technology; or circus skills, street arts or other physical theatre activity

iv A historic park or garden open to the public; A historic place of worship attended as a visitor (not to worship); A monument such as a castle, fort, or ruin; or a site of archaeological interest

